# Non-pharmacological interventions for psychotic symptoms in people with dementia: a Delphi consensus

**DOI:** 10.1101/2024.09.12.24313578

**Authors:** A Choi, J McDermid, K Mills, A Sweetnam, J Fossey, Z Khan, C Ballard

## Abstract

**Background:** Psychotic symptoms such as delusion and hallucinations are common in people with dementia. They are associated with various deleterious outcomes including reduced quality of life and increased caregiver burden. Pharmacological interventions to combat psychotic symptoms have shown limited efficacy and can be associated with significant adverse events. Non-pharmacological interventions are recommended as the first line option for treatment, however there is a paucity of evidence for specific non-pharmacological options to primarily target psychotic symptoms in people with dementia. Further work is needed to identify, adapt and develop possible non-pharmacological options to target psychotic symptoms in dementia.

**Aim:** To establish which non-pharmacological interventions could be used or adapted to treat psychotic symptoms and/or confer benefits in people with dementia

**Design:** Modified Delphi consensus process. Two rounds of feedback were conducted and included a directed scoping review, based on the interventions recommended in the first round of the Delphi.

**Participants:** An expert panel consisted of 12 members with clinical and research expertise in managing psychotic symptoms in people with dementia

**Results:** There were three top nominated treatment options: cognitive behavioural therapy (CBT), family intervention, and personalized activities/environmental/sensory interventions, without a clear priority between the 3 approaches.

The WHELD/Brief Psychosocial Therapy programme focussing on personalized activities improves concurrent neuropsychiatric symptoms in people with dementia related psychosis. Preliminary studies also suggest that combining personalized activities with family training may improve the direct impact on psychosis. There are also opportunities to adapt CBT interventions for people with psychosis related to early or mild dementia.

**Conclusions:** There were clear recommendations for three non-pharmacological options that could be used or adapted to benefit people with psychosis in the context of dementia.

## Introduction

Dementia is often considered to be a disorder of cognition and function, associated with difficulties in everyday living. However, neuropsychiatric symptoms are almost universal and frequently include psychotic symptoms (Ballard, Saad, et al., 1995). Psychotic symptoms are associated with reduced QoL and wellbeing in people with dementia living in the community and nursing homes, respectively. Furthermore, the association between psychotic symptoms and reduced QoL is mediated by co-morbid neuropsychiatric symptoms, such as agitation, aggression and depression.

There is a paucity of non-pharmacological treatment options targeting psychotic symptoms in people with dementia. Guidelines encourage the use of non-pharmacological interventions as the preferred initial approach to manage or treat psychotic symptoms. Non-pharmacological approaches have been shown to be effective for agitation/aggression, apathy and depression, but the choice of intervention is crucial to obtain the most beneficial outcomes (Testad et al., 2014). Importantly however, in a Delphi consensus conducted in 2019, there was insufficient evidence to recommend any specific non-pharmacological treatment for psychosis in people with dementia, (Ballard, Kales, et al., 2020; Kales et al., 2019).

Given the limited literature specifically evaluating the impact of non-pharmacological interventions on symptoms of psychosis in people with dementia is limited, an approach is needed to consider both the available information and potential opportunities to inform the development or adaptation of other non-pharmacological interventions which could give benefit.

The present study uses a modified Delphi consensus to identify possible candidate non-pharmacological interventions for psychosis in people with dementia and to identify other interventions which could be adapted as treatments for specific psychotic symptoms in people with dementia.

### What is a Delphi?

A Delphi is a structured means of opinion gathering from groups of experts, in order to solve a complex problem or create novel ideas. It achieves this goal through several iterations of questionnaires and tailored feedback in order to filter opinions and achieve consensus (Day & Bobeva, 2005). Named after the oracle at Delphi, the technique traces its origins to the Rand Corporation, the original purpose of which was to make technological forecasting in the defence industry (Dalkey & Helmer, 1963). Whilst the methodology has gone through various changes, two fundamental characteristics remain from the approach that was first introduced by Dalkey and Helmer (1963). 1) The use of iterations or “rounds”, where each round of questions presented to experts reflect modifications based on the input of the experts from previous rounds. 2) The results of each round are fed back to the experts, allowing them to change their view if necessary (Barrett & Heale, 2020).

A previous Delphi study has been conducted to establish appropriate treatment for neuropsychiatric symptoms in Alzheimer’s disease (Kales et al., 2019). The Delphi was able to identify and recommend non-pharmacological approaches for agitation and overall neuropsychiatric symptoms, encouraging the use of adaptation of the environment, person-centred care, tailored activities, and caregiver training. Interestingly however, given the limited evidence, the consensus panel felt unable to make specific recommendations regarding non-pharmacological treatments for dementia related psychosis.

The findings, presented by Kales et al. (2019), imply that a broader question with fewer restrictions needs to be proposed to experts regarding non-pharmacological treatments for psychotic symptoms in dementia. The question needs to include the potential to adapt and develop non-pharmacological interventions, in addition to considering evidence that is already available.

The findings from Kales et al. (2019) also imply that any evidence needs to be gathered and made available to inform the expert opinions and help the experts to make more consistent recommendations. This can be achieved through a modified version of the Delphi, where a scoping review is initially directed by suggestions from the expert panel in the first iteration, then included within the feedback process as part of the next iteration. This use of a Delphi, with the inclusion of a scoping review as a feedback option, is not new and has been used previously to identify treatment directions for drug repurposing in Alzheimer’s disease (Ballard, Aarsland, et al., 2020; Corbett, Pickett, et al., 2012).

## Methods

### Study Design

A Delphi design was utilized to capture and aggregate opinions and achieve consensus from a panel of experts in order to establish which non-pharmacological treatments may be most suitably used or adapted to treat or benefit people with dementia related psychosis. The Delphi process involved two rounds of anonymized opinion gathering, where the feedback was summarized and fed back to expert panel members together with a scoping review of the top three non-pharmacological treatment options identified in Round 1, the results of which were provided to expert participants as part of the feedback to inform their considerations in Round 2 (following the method of Corbett et al., 2012). This Delphi study was reviewed and approved by the University of Exeter college of medicine and health research ethics committee (Ref: Sept20/D/251).

### Expert panel participants

There were 14 expert panel members from a variety of geographical areas included in Round 1. Two panel members did not continue to Round 2. The 12-expert panel members that completed the study are listed below. Each expert panel member was required to have:

1. A relevant profession, such as a medical practitioner, psychologist, care provider or researcher with expertise in symptom treatment and management of psychotic symptoms in older adults or psychological therapies for psychosis in other populations.
2. Published research relevant to dementia and/or therapies for psychotic symptoms.

List of Experts: Clive Ballard, Linda Clare, Esme Moniz Cook, Jan Oyebode, Daniel Collerton, Reinhard Gus, Zunera Khan, Bob Woods, Elizabeth Kuipers, David Kingdon, Dag Aarsland, and Jane Fossey

### Delphi consensus criteria

For this study, consensus was defined as percentage agreement across the expert panel members following the second round of prioritisation. The criteria for agreement were 75% in order to ratify consensus.

### Delphi round 1 method

During the first round of the Delphi process expert panel members were contacted by email and asked to complete an online survey. They were asked to suggest 3 to 5 non-pharmacological therapies or therapeutic strategies addressing psychotic symptoms that they thought could readily be adapted to treating or managing psychotic symptoms in people with mild-to-moderate dementia. They were then asked to prioritise their listed therapies/strategies and explain the reason for their selection and prioritization. The suggestions were compiled, ranking each non-pharmacological treatment according to the number of times it was suggested. A scoping review was then undertaken for each of the top three suggestions, to identify existing evidence supporting these approaches in relation to the treatment of psychotic symptoms in people with dementia

#### Directed scoping review method

A scoping review was then used to explore the available evidence (Munn et al., 2018) for the top three non-pharmacological treatment options proposed by the panel. (Table 1).

**Table 1:** Summary of the panels top nonpharmacological suggestions in Round 1.

| **Intervention Listed** | **Panel members recommends** |
| --- | --- |
| Cognitive Behavioural Therapy (CBT) | 7 |
| Family Intervention/Education | 4 |
| Activity/Environment/Sensory Intervention | 4 |

The search criteria were as follows:

> The primary aim of the scoping review was to detect studies related to each of the three treatment approaches suggested by the panel as possible treatments for psychotic symptoms in people with dementia.
>
> Databases: Pubmed, Web of Science, EBSCOhost
>
> Search strings were developed for the following key concepts:
>
> 1. Dementia: e.g. [dementia OR Alzheimer*]
> 2. Psychotic symptoms: e.g. [delusion* OR hallucination* OR psychosis OR psychotic*]
> 3. Non-pharmacological interventions: e.g. [treatment* OR therap* OR counsel* OR intervention* OR non-pharmacologic*]
> 4. Specific strings were developed to focus on the top three interventions suggested by the panel
>   a. Cognitive Behavioural Therapy: e.g. [“cognitive behavioural” OR CBT]
>   b. Family Intervention/Education: e.g. [famil*]
>   c. Activity/Environment/Sensory Interventions: e.g. “Activity” or “personalized activity” OR “reminiscence” OR “music” OR aromatherapy [“environment*” OR “room design” OR “perceptual changes” OR “bright light therapy” OR “proprioceptive sensory input” OR “functional task object” OR “garden” OR “black tape grid” OR “sensory devices” OR “tinted lens”]
>
> As the goal was to be as inclusive as possible, RCTs, Quasi experimental studies, observational studies and case series were included.

### Delphi round 2 method

The three recommended non-pharmacological approaches from round 1 were scored in priority order by each of the Delphi panel, and their prioritizations were counted and provided to the panel along with the results of the directed scoping review. The panel were then asked to consider 2 options:

a. Recommend using one of the interventions identified by the scoping review, as a treatment for psychotic symptoms in people with dementia.
b. Focus on building a new approach and adapting one or more of the top 3 interventions recommended by the panel.

### Results

#### Round 1

Round 1 of the Delphi generated 22 suggestions for different non-pharmacological treatments. The treatments with the most nominations are summarised in Table 1.

The three most nominated interventions were cognitive behavioural therapy, family intervention/ education and activity/environmental/ sensory interventions. The latter group was a diverse series of specific suggestions pulled together under a broad heading. A scoping review was conducted to identify published evidence pertaining to these three interventions.

In the scoping review, the primary outcome measure for most of the identified studies was either overall neuropsychiatric symptoms, but psychotic symptoms in dementia were reported as a secondary outcome measure in some studies.

The process for identifying and screening articles for inclusion in the scoping review is shown below in Figure 1.

**Figure 1.**
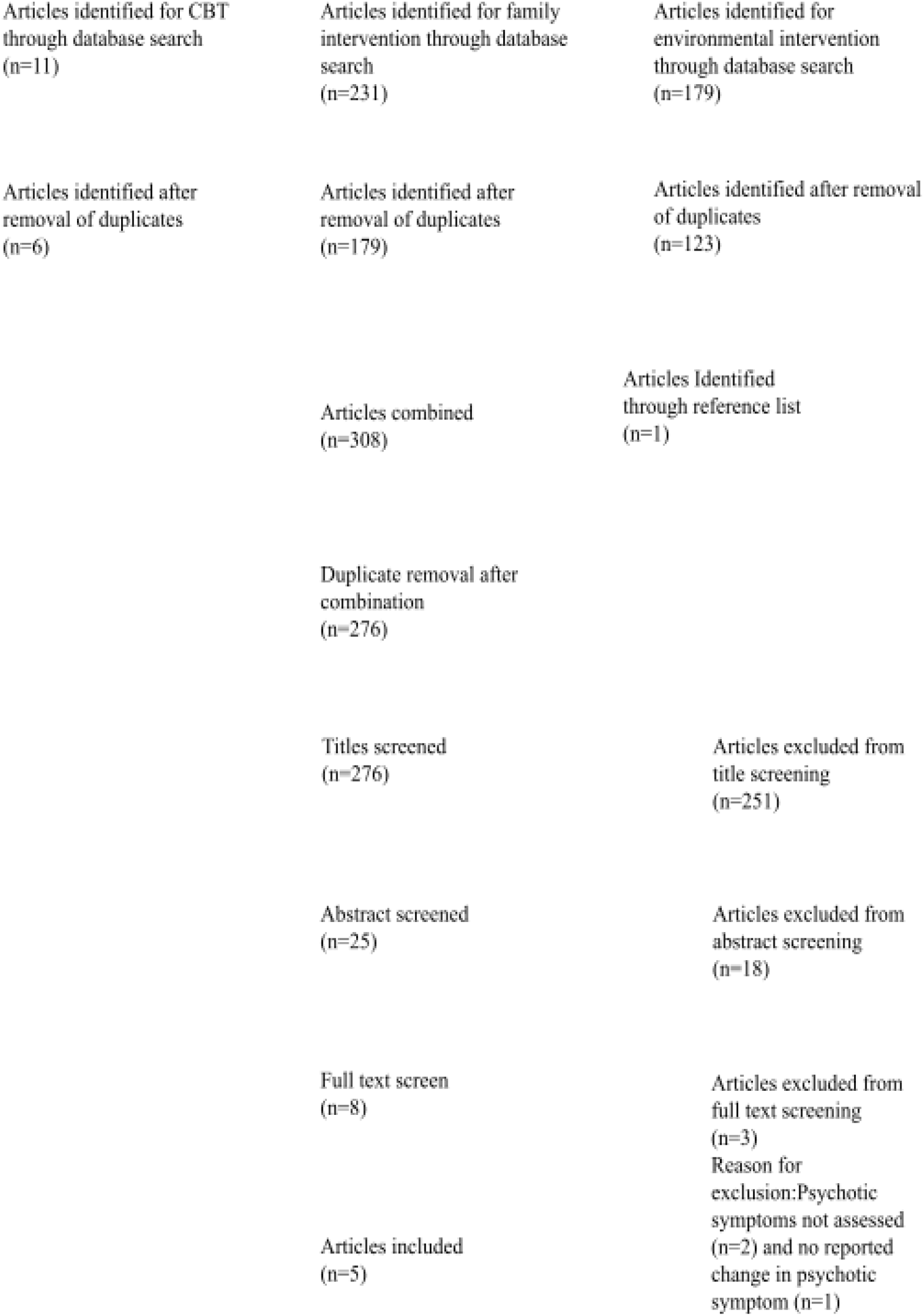
Screening process for article inclusion in scoping review.

A summary of the results from the articles identified in the scoping review is shown below in Table 2, and the studies are then described in more detail in the subsequent section.

**Table 2:** Results of the directed scoping review nonpharmacological interventions focusing on psychotic symptoms in people with dementia.

| **Study/authors** | **Design** | **Intervention** | **Number of participants** | **Duration** | **Assessment Tool** | **Main findings** | **Cochrane quality rating (RoB-2)** | **Panel suggestion mapped** |
| --- | --- | --- | --- | --- | --- | --- | --- | --- |
| Chen et al. (2014) | Cohort Study with reference group | Organized mixed program | 104 veterans home residents | 12 weeks | NPI | Reduced NPI scored hallucinations ($-0.82$, $p = 0.004$) and delusions ($-0.9$, $p = 0.018$) | n/a | Activity/Environment/Sensory Intervention |
| De Oliveira et al. (2019) | RCT | Tailored Occupational Therapy | 21 Outpatients and carers | 3 Months | NPI-C | Reduced NPI scored hallucinations in treatment group ($-1.25$, $p = 0.04$) | Low concern | Family Interventions |
| Nobili et al.<br>(2004) | Pilot-R<br>CT | Family<br>Intervention | 69<br>Outpatients<br>and carers | 1<br>session,<br>6 & 12<br>month<br>follow-up | Spontaneous Behavior<br>Interview<br>(SBI-C) | Reduced<br>delusions in<br>treatment<br>group($p=0.04$) | Some<br>Concern | Family<br>Interventions |
| Schindler et<br>al. (2002) | Case<br>Study | Bright Light<br>Therapy | 5 patients | 14 days | Confusion<br>rating<br>scale(CRS) | Reduced<br>delusions in 3<br>patients,<br>Increased in<br>psychotic<br>symptoms in 1<br>patient | n/a | Activity/Environment/Sensory<br>Interventions |
| Ballard et al<br>2018, 2024 | Factorial<br>RCT | Brief Psychosocial Therapy<br>module of<br>WHELD<br>programme | 163 | 9<br>months | NPI<br>psychosis<br>score<br>NPI apathy<br>CMAI | No improvement in<br>psychosis, but<br>significant<br>improvements<br>in agitation and<br>apathy in<br>people with<br>psychosis | No<br>concern | Activity/Environment/Sensory<br>Interventions |

#### Nonpharmacological interventions identified by the scoping review

##### Activity/Environmental/Sensory Intervention identified by scoping review

Ballard et al (2018) conducted a cluster RCT in 847 people with dementia in 69 UK nursing homes. Participating nursing homes were randomized to receive either the WHELD intervention (combining person centred care training, antipsychotic review, Brief Psychosocial Therapy (personalized activities or exercise with social interaction) or treatment as usual in a 1:1 ratio. In the original study there was a significant reduction in agitation in the participants receiving WHELD compared to those receiving treatment as usual, but there were no direct benefits in psychosis. A subsequent paper (Ballard et al 2024), has reported an additional exploratory analysis of the 163 patients who participated in the study who had psychosis at baseline (a score of 1 or more combining the NPI scores for hallucinations and delusions). There were no differences between groups in the NPI psychosis scores, but in the patients with psychosis, the WHELD/Brief Psychosocial Therapy intervention conferred significant benefits in agitation (CMAI p=0.035) and apathy (NPI apathy domain p=0.006) and a significant benefit in quality of life (DEMQOL proxy p=0.01) compared to the treatment as usual group..

Chen et al. (2014) utilised an organised mixed program consisting of music therapy, orientation training, art-cognitive activities, physical activities and staff education. The 104 participants were split into two groups: experimental versus reference group, but there was no blinding or random allocation. The reference group only received staff education. Medication was standardised between the groups. The primary research outcome was to investigate changes in overall neuropsychiatric symptoms. The research was able to identify significant improvement in overall neuropsychiatric symptoms using the NPI. As secondary outcomes, using the individual subcategories from the NPI, the study reported improvements in both the delusions and hallucinations domains in the experimental group after 12 weeks…

Schindler et al. (2002) conducted bright light therapy to evaluate potential improvements in delirium or confusional states, which may include psychotic symptoms. The therapy was conducted on 5 participants with dementia. It involved exposing individuals to bright light, two hours a day for 14 consecutive days. Three participants showed reduced psychotic symptoms, it should be noted that one participant had no prior psychotic symptom. Interestingly, one participant showed increased symptoms of delusions and agitation. This participant developed blurred vision after 8 days and signs of hallucinations after exposure to bright light for 12 days. Therapy for this individual was discontinued and their delusions and hallucinations stopped after one day.

##### Family Interventions identified by scoping review

De Oliveira et al. (2019) adapted the Tailored Occupational Therapy (TAP), developed by Gitlin et al. (2009), to targeting outpatients receiving treatment from memory clinics. This specific adaptation was called Tailored Occupation Therapy-Outpatient (TAP-O). The program involves an occupational therapist developing activities based on capabilities, interests and roles of people with dementia and adapting these activities to their cognitive and functional ability. The aim was to identify changes in severity of any neuropsychiatric symptoms included within the 12 NPI domains, including delusions and hallucinations. The study was randomiized, but was a small pilot with 21 participants and did not correct for multiple endpoints. The study reported a significant reduction in the severity of hallucinations (*p*=.04) but not delusions over the 3-month intervention.

Nobili et al. (2004) evaluated the effect of a structured family intervention. The intervention consisted of separate home visits by, 1) a psychologist for 60 minutes and 2) an occupational therapist for 90 minutes. The psychologist reviewed and discussed family dynamics, caregiver stress, communication techniques and techniques to manage challenging behaviours experienced by people with dementia. The occupational therapist provided practical information on controlling reactive behaviour, modifying retained functional abilities and altering the home environment to reduce the risk of harmful situations. The intervention was compared to a group that received support through a helpline made available to families which provided information about practical issues such as financial support and community services/clinics. The primary outcome measures were caregiver stress, the rate of institutionalisation of people with dementia and the severity of challenging behaviours (which included motor agitation, hallucinations, and delusions. At a 12 month follow up, the study reported a reduction in overall scores for challenging behaviour scores (*p*<0.03) and a reduction in the amount of care time needed for the intervention group. As a secondary outcome, the authors found a significant decrease in the frequency of delusions for the intervention group.

#### Strength and limitations of the identified nonpharmacological interventions

To rate the quality of studies identified, the Cochrane risk of bias tool (ROB 2) (Cumpston et al., 2019) was used. Three studies randomized participants, meeting the required prerequisite in order to be rated by the ROB 2,. The studies conducted by De Oliveira et al. (2019) and Ballard et al (2018) showed ‘Low concern” as they utilized reported adequate randomisation process and concealment The study conducted by Nobili et al. (2004) was rated as “Some concern” using the Cochrane framework. Whilst the study reported adequate randomisation measures and good concealment, there were high drop-out rates which differed between treatment groups.

The two other studies identified from the scoping review, were not randomised and hence did meet the criteria to be rated according to the ROB 2. The study conducted by Chen et al. (2014) compared outcomes to a reference group, limiting interpretation of results. In respect to the Schindler et al. (2002) study, only 5 participants were reported and findings of the study were mixed in terms of treatment effect.

### Round 2

Out of the 14-panel members who initially responded, 12 members continued on to Round 2, and were asked to nominate their top intervention that would most readily be developed or adapted. A choice was given to the panel whether a new intervention should be developed from the nominations of round 1 or an intervention identified from the scoping review should be adapted and refined for the specific purpose of targeting psychotic symptoms in dementia. There were 4 experts who nominated CBT, 5 experts nominating family intervention or education, and 2 nominated activity, environmental or sensory interventions. One individual opted for a specific intervention identified by the scoping review namely; the TAP-O intervention reported by De Oliveira et al. (2019).

The Delphi did not achieve consensus and did not proceed to a following iteration for two reasons 1) Diverging opinions meant that reaching a 75% consensus threshold was unlikely and 2) Most experts recommended a combination of non-pharmacological interventions to accommodate the wide range of individual differences contributing to psychotic symptoms in people with dementia.

## Discussion

The present chapter reports a Delphi consensus study to establish which non-pharmacological treatments are most suitable for use or adaptation into an intervention to treat or manage psychotic symptoms in people with dementia.

The top nominations for non-pharmacological treatment included CBT, family interventions and activity/sensory/environmental intervention. The Chen and D’Oliveira studies both combined elements of enjoyable/personalized activities and family training, the WHELD/Brief Psychosocial Therapy programme combined personalized activities with training of professional Care staff.

Based on the current evidence, the only substantial RCT to examine a non-pharmacological approach with psychosis as an outcome is the WHELD/Brief Psychosocial Therapy programme, which did not show direct benefits in the treatment of psychotic symptoms, but did improve outcomes for people with dementia experiencing psychotic symptoms by improving concurrent neuropsychiatric symptoms and quality of life.

The Chen study combined personalized activities, mainly music, with family training; and demonstrated benefits in psychosis as a secondary outcome. However, the study was not randomized and the results need to be interpreted cautiously. The D’Oliveira study also combined a personalized activity approach with family training in a small (n=21) pilot RCT, which did show benefits in psychotic symptoms in the main analysis which did not control for multiple outcomes. Both studies are therefore promising, but need larger studies to confirm the benefits. They do also highlight the potential benefits of combining personalized activities with family training, which may also be a further opportunity to optimize the WHELD/Brief Psychosocial Therapy Intervention.

The panellists who opted for CBT, focussed on interpretation of stressors leading to psychotic symptoms and the management of stress and anxiety. : nCBT could be adapted to target psychotic symptoms, but would need to be simplified to suit individual cognitive abilities and to incorporate elements of trigger monitoring specific to psychotic symptoms; but this approach is probably more suited to people with psychosis in the context of early or mild dementia.

In summary, there is evidence that the WHELD/Brief Psychosocial Therapy programme, focussing on promoting personalized activities, improves concurrent neuropsychiatric symptoms in people with dementia related psychosis. Preliminary studies also suggest that combining personalized activities with family training may improve the direct impact on psychosis. There are also opportunities to adapt CBT interventions for people with psychosis related to early or mild dementia.

## Data Availability

All data produced in the present study are available upon reasonable request to the authors

